# Black Summer bushfires and psychological distress of young people: what mental health, economic, social, and community resources are risk and protective factors?

**DOI:** 10.1101/2025.06.25.25330259

**Authors:** Ben Edwards, Matthew Gray

## Abstract

**Objectives:** To systematically assess whether economic, social and community resources can protect against the impact of the Australian “ Black Summer” bushfires on adolescent psychological distress. Using 16 years of longitudinal data, we also test whether history of mental health problems, history of economic hardship puts youth at risk and whether a history of high parental warmth and low harsh parenting protects young people.

**Study design:** Prospective, population-based cohort study; analysis of Longitudinal Study of Australian Children (LSAC) survey data

**Setting, participants:** Adolescents in the nationally representative cross-sequential sample of Australian children recruited in 2004 for the Birth and Kindergarten cohort (aged 0-1 and 4-5 years at enrolment). Survey data from waves 9 (16-17 and 18-19 years for B and K Cohort) and waves 1 to 8

**Main outcome measures:** Psychological distress measured by the Kessler 10-item questionnaire, K10.

**Results:** There were 2,726 respondents from the B and K Cohorts who had psychological distress information and demographic characteristics in Wave 9. We did not find that a history of financial hardship, unemployment, being partnered, high social support, household harmony and additional stressful life events moderated the association between fires and psychological distress. Family financial hardship or capacity to raise $2000 in an emergency in the previous wave, or over the prior 16 years did not increase or reduce the risk. Community collective efficacy, and history of positive parenting (high warmth, low harshness) was also not significant. History of high anxiety but not depressed mood was the only significant risk or protective factor. Those with a significant history of high anxiety had elevated distress levels when exposed to fire in the last 12 months compared to those with none or some history of high anxiety.

**Conclusions:** Additional targeted support for youth for those with prior history of anxiety through the Better Access initiative may be particularly important. However, our findings do underscore that leaving prior mental health history aside, how robust the impacts of bushfires were on youth mental health and that being exposed to the Black Summer bushfires affected all youth irrespective of privilege or social address.

## Introduction

Studies of adults suggest that there were mental health impacts of the Black Summer bushfires with higher depression, anxiety and stress reported by those affected compared to those with lower levels of exposure or elevated clinical cut offs (MacLeod et al., 2023, Usher et al., 2022). There is limited evidence on the impacts of Black Summer on children and adolescents with only one large, but not representative study of adolescents suggesting that those exposed to fires had greater risks for suicidal ideation, trauma and insomnia (Beames, J.R., Huckvale, K., Fujimoto, H. *et al* 2023). Recent longitudinal research in Australia suggests that fires or floods were associated with increased risk of self-harm and suicidal ideation in adolescents 14-19 years however for younger children single disaster exposures such as bushfires were not associated with mental health (Edwards, Taylor & Gray, 2024; Campbell, Edwards & Gray, 2024).

A recent scoping review focussed on identifying risk and protective factors in response to climate change-related natural disasters (Ma, Moore & Cleary, 2022). While only eight studies examined fires, overall they reported that protective factors included family factors as positive parenting styles and parental relationships, social support (particularly emotional support), positive coping strategies, and community connection and volunteering. Risk factors included negative parenting styles, additional stressful life events, and community violence. Of the 92 articles identified in the review only 28% were longitudinal studies, with only 7% having baseline mental health assessments. A meta-analysis of mental health consequences beyond 12 months following disaster exposure of 234 papers rerpoarted that depression and anxiety remained elevanted and was significantly higher for children and adolescents (Newnham et al., 2022). While 75% of the risk and protective factors tested in these studies were not significant, risk factors found to be associated with higher levels of anxiety and depression included a history of psychiatric/anxiety/depression, negative life events post-disaster, and poor social ties. Protective factors included having a partner, fewer subsequent life stressors and high social support. There were only three studies of fires in the meta-analysis (Newnham et al, 2022) while the scoping review (Ma et al., 2022) included just eight studies with only one longitudinal study of adolescents (Langley, & Jones, 2005), suggesting that more research on child and youth risk and protective factors are needed.

While the role of risk and protective factors for mental health problems has been the focus of much adult research, few studies have focussed on factors that make youth vulnerable or resilient to mental health problems following bushfires and even fewer examined this longitudinally. The Black Summer bushfires had a particularly large scale and scope nationally and internationally. This paper systematically tests whether economic, social and community resources can protect against any of the adverse effects of fires on the mental health of youth. Using the national cohort study of children, it is uniquely placed to test whether history of mental health problems puts young people at greater risk across a large national sample. This longitudinal data also affords the opportunity to test the longer-term history of economic resources and parenting history affords any additional protection.

## Method

### Study design and participants

This study primarily used wave 9 of the Longitudinal Study of Australian Children (LSAC) although variables from waves 1 to 8 were also used. LSAC, also known as *Growing Up in Australia*, is a large, longitudinal, nationally representative cohort study of Australian children. The survey has been administered every two years since 2004 (wave 1), with wave 9 administered in 2020-2021. The survey has followed two cohorts, the B-Cohort (birth cohort, born in the year preceding wave 1), aged 16-17 in wave 9, and the K-cohort (kindergarten cohort, aged 4-5 in wave 1), aged 20-21 in wave 9 (Growing up in Australia, 2021).

Wave 9 C1 was in field from October-December 2020 (ADA, 2022) and captured exposure to the Australian “ Black Summer” bushfires of 2019-20. Waves 1 through 8 consisted of in-person interviews with study children (from 10 years of age) and primary caregivers (from wave 1). However, the Australian Bureau of Statistics interviewers were not able to conduct face-to-face interviews due to the pandemic. Instead, an online survey was administered to young people and their parents and there was a shorter fieldwork period than the six-month period. Wave 9C1 consisted of 2,017 respondents from the B-cohort, and 1,786 respondents from the K-cohort.

Disaster exposure from waves 4 to 8 was used as a control variable in early analysis.

Results are given in the Appendix.

LSAC is a sample is nationally representative of children within each cohort drawn using Australia’s public healthcare (Medicare) enrolment database. In 2004, the Medicare dataset covered virtually all four-year old children and 88.5% of children under 12-months (Soloff, Lawrence & Johnstone, 2005). Cross-sectional sample weights, provided by the data custodians to address non-response error, were used in analysis.

The Australian Institute of Family Studies Ethics Committee provided ethics approval for the LSAC, and all participants provided written informed consent.

### Dependent variables

*Kessler 10-item questionnaire (K10)*. Developed by Kessler and others to use in the US National Health Interview Survey measure psychological distress (Kessler, Andrews, Colpe et al., 2002), the K10 has been widely used in screening in population health surveys in Australia (Slade, Grove & Burgess, 2011).

### Disaster exposure variables

The LSAC has collected data on exposure to natural disasters since wave 4. In wave 9, the survey asked adolescents “ Have you been affected by any of the following extreme weather events or natural disasters in the past 12 months? Bushfire; Drought; Flood; Storm/Hail; Cyclone; Other extreme weather events or natural disasters” . The survey yields six corresponding self-report measures: one for each type of disaster. In waves 4 to 8 primary caregivers’ reported on whether they have been affected by natural disasters in the previous year. In these waves, caregivers reported on whether in the last 12 months caregivers “ had your home or local area affected by bushfire, flooding or a severe storm” , and whether in the past 12 months the caregiver “ lived in a drought-affected area” .

### Risk and protective factors

#### Demographic characteristics (Wave 9)

Statistical models included child and family/neighbourhood characteristics as covariates and tested for interactions with fire exposure. Child characteristics were: sex, and the cohort to which the child belongs (B-cohort: aged 16-17; or K-cohort: aged 20-21).

Family/neighbourhood characteristics included whether the family had moved house since the last wave, and whether they lived in a regional or metropolitan area.

#### Economic resources

Economic Hardship was measured using a 6-item scale of financial hardship (e.g. “ Over the last 12 months, due to shortage of money, have… you not been able to pay gas, electricity or telephone bills on time” ?). Questions capture whether a household has struggled to meet basic living expenses such as food, rent and utilities in the past 12 months. For Wave 9 the respondent was the Young Person, and we coded financial hardship as 0 for no hardships, 1 for one hardship and 2 for two or more hardships. For history of financial hardship, the primary caregiver reports of hardship was used, with the proportion of waves where the Young Person was in a household where there were 2 or more hardships calculated. This was coded as 1 for no history at all, 2 for more than 0 but less than 0.40 waves with two or more hardships and 3 if there were 0.40 or more waves with two or more hardships.

*Employed* measures whether the Young Person was employed in any capacity. *Raise $2000 in an emergency* was asked to primary caregivers from waves 1 to 8 with the following: “ Suppose you only had one week to raise $2000 for an emergency. Which of the following best describes how hard it would be for you to get that money? “ Primary caregivers could indicate that they “ Could easily raise the money” , “ Could easily raise the money but it would involve some sacrifices (e.g. reduced spending, selling a possession), “ You would have to do something drastic to raise the money (e.g. selling an important possession)” , “ You don’t think you could raise the money” . For Wave 8 we coded “ easily” as 1, “ sacrifices” as 2, “ drastic” and “ don’t think you could” as 3. For history of difficulties raising money, we calculated the mean of the 1 to 3 for all waves that primary caregivers participated in. Any scores above 2 were then coded as 1, and those below 2 was coded as 0.

#### Social resources of young people

*Partnered* indicated whether the Young Person was partnered (1 = yes, 0 = no) for Wave 9. *Social support* was measured using an adapted 15-item version of the Medical Outcomes Study Social support scale which captures emotional/informational, tangible, affectionate support as well as positive social interaction (Ware, Nelson, Sherbourne & Stewart, 1992). *Get along with others in the household*. In Wave 9 Young People were asked “ Sometimes household members may have difficulty getting along with one another. They do not always agree and they may get angry. In general, how would you rate your household’s ability to get along with one another?” Young people provided ratings from “ Excellent” to “ Poor” . Responses that indicated “ fair” or “ poor” were coded as 1, while all other responses (Excellent, very good, good) were coded as 0.

#### Parenting history: High parental warmth and harsh parenting

*Parental warmth* was measured with a 6-item scale (e.g. “ How often do you… feel close to this child both when he/she is happy and when he/she is upset), capturing how often a parent expresses affection, nurtures and supports a child. Primary caregivers commonly reported high scores for warmth, so for average item scores of 4 out of 5 or more over all the waves, we coded this as 1 and for those that were under 4 were coded as 0.

*Harsh parenting* was measured using adapted items from the Early Childhood Longitudinal Study of Children, Birth Cohort and the National Longitudinal Survey of Children and Youth 1998−1999 for Waves 1 to 2. For waves 3 to 6: Items were adapted from the Ineffective/harsh Parenting scale developed for the National Longitudinal Study of Children and Youth (NLSCY). First we coded scores in the top 25% across the eight available waves as 1 and less as 0 for each wave (2.31 being the cut-off). Next we pooled these scores over the eight waves, and then calculated tertiles for this recoded harsh parenting measure. Because many primary caregivers reported very low scores, the first tertile accounted for 48.6% of young people’s parenting experiences while the second tertile accounted for 25.1% and the final tertile was 26.3%.

#### History of mental health problems

*Depression* was measured in LSAC at ages 12–13 and 14–15 and 16–17 using the 13-item Short Mood and Feelings Questionnaire (Angold, et al., 1995). Scores ranged from 0–26 with higher scores reflecting a greater level of depression. Clinical cut-offs for elevated depressive symptoms were indicated for a sum score of 8 or higher while scores lower than 8 are indicative of low depressive symptoms (scored one and zero, respectively). For history of depression we calculated the mean of clinical cut offs for every wave participated. This was then coded as 0 for no history of clinical levels over any waves, 1 if less than half the waves there was a history of clinical depression and 2 if over half the waves there was a history of clinical levels of depression.

*Anxiety* was measured at ages 12–13 and 14–15 and 16–17 using the 8-item Spence Anxiety Scale, short form (Spence, 1998). Scores ranged from 0–24, with higher scores reflecting higher levels of anxiety symptoms. At each age, anxiety scores above 8 (top 20%) were indicative of elevated anxiety symptoms and coded 1. Lower scores were coded 0. For history of high anxiety we calculated the mean of every wave participated and then coded this as 0 for no history of high levels over any waves, 1 if less than half the waves there was a history of high levels of anxiety and 2 if over half the waves there were high levels of anxiety.

### Other covariates

Statistical models also included child and family/neighbourhood characteristics as additional covariates. Child characteristics were: child sex at birth, whether the child was born overseas, and whether the child is Aboriginal or Torres Strait Islander.

Family/neighbourhood characteristics included their state of residence. Given that policy restrictions and COVID-19 outbreaks varied substantially by states and territories, regional and state of residence dummies also serve to control for variation in experiences in the pandemic (Edwards et al., 2022). Neighbourhood socioeconomic characteristics were measured using Socio-Economic Indexes for Areas (SEIFA) scores (ABS, 2022). We used deciles of the SEIFA Index of Relative Socioeconomic Advantage/Disadvantage, a summary measure of the social and economic conditions of Australian neighborhoods. We also included exposure to disasters in the prior decade as a covariate. This also took account the long-term impacts of disasters so that the mental health impacts of Black Summer and other disaster exposures in the prior 12-month period are identified.

### Statistical analyses

Several of linear regression models predicting psychological distress were estimated. Model 1 assessed the impact of exposure to bushfires and interaction with other demographic characteristics on psychological distress. Model 2 included the same covariates as model 1 but tested statistical interactions between fires and economic resources at Wave 9 (employment and hardship). Model 3 added social resources to Model 2 (partnered, social support, get along others in household) and tested interactions between fire and these variables. Model 4 added stressful life events and tested the interaction with fire on distress. Model 5 tests the interaction of economic resources from wave 8 only (hardship and raise $2,000 in an emergency) with fire on distress and included all Model 1 covariates. Model 6 included Model 1 variables and then separately tested interactions between fire and history of mental health problems (depression and anxiety), economic resources (hardship and raise $2,000 in an emergency), community resources (collective efficacy) and parenting (high warmth, low harsh) on distress in wave 9.

All statistical interactions were tested with F tests and significant interactions displayed in figures with 95% confidence intervals.

All models were run with the provided cross-sectional sample weights, which correct for survey non-response using Stata 15.

### Ethics approval

The Longitudinal Study of Australian Children was approved by the Australian Institute of Family Studies Ethics Committee.

## Results

In total there were 2,726 respondents who had psychological distress information and demographic characteristics. Table 1 shows rates of exposure to natural disasters, along with impacts reported for those exposed, along with demographic and geographic characteristics of the weighted sample. The table shows that 10.2% of adolescents were exposed to bushfires in the 12-months prior to the survey, and 8.8% were exposed to a storm or cyclone. 18.3% were exposed to at least one disaster, 6.8% exposed to more than one disaster, and 27.7% had previously identified a disaster exposure (in waves 4 to 8).

**Table 1:** Demographic, geographic characteristics and disaster exposure rates, weighted percentage.

| Variable | Weighted percent |
| --- | --- |
| *Geography* |  |
| Lives in urban area | 70.02 |
| *State* |  |
| NSW | 31.54 |
| Vic | 25.92 |
| Qld | 21.37 |
| SA | 6.34 |
| WA | 9.30 |
| Tas | 2.91 |
| NT | 0.43 |
| ACT | 2.19 |
| *SEIFA decile* |  |
| 1 | 6.53 |
| 2 | 8.23 |
| 3 | 10.48 |
| 4 | 8.94 |
| 5 | 11.66 |
| 6 | 10.28 |
| 7 | 11.30 |
| 8 | 9.92 |
| 9 | 10.85 |
| 10 | 11.81 |
| *Demographic characteristics* |  |
| Female | 49.35 |
| *Cohort (age)* |  |
| Birth cohort (16-17 yrs) | 53.02 |
| Kindergarten cohort (20-21 yrs) | 46.98 |
| Indigenous | 2.20 |
| Born overseas | 2.13 |
| Moved in last wave | 11.01 |
| *Disaster exposure* |  |
| Exposed to bushfire | 10.22 |
| Exposed to drought | 6.16 |
| Exposed to flood | 2.13 |
| Exposed to storm/cyclone | 8.80 |
| Exposed to other disaster | 0.54 |
| Previously exposed to any disaster (w4 to 8) | 27.70 |

Table 2 shows the significant interactions tested and that there was a significant interaction (at the p<.10 level) between fire exposure at Wave 9 and history of high anxiety on psychological distress. No other interactions were significant.

**Table 2:** Statistical interactions between bushfire in the past 12 months and risk/protective factors, weighted linear regression.

| Risk/protective factors | F | df | Significance | N |
| --- | --- | --- | --- | --- |
| **Wave 9 factors** |  |  |  |  |
| *Demographic characteristics<sup>1</sup>* |  |  |  |  |
| Sex | 0.37 | 1 | 0.543 | 2726 |
| Cohort | 0.47 | 1 | 0.492 | 2726 |
| Urban | 0.04 | 1 | 0.833 | 2726 |
| Moved in last wave | 0.09 | 1 | 0.765 | 2726 |
| *Economic resources<sup>2</sup>* |  |  |  |  |
| Hardship | 1.07 | 2 | 0.343 | 2712 |
| Employed | 0.02 | 1 | 0.878 | 2712 |
| *Social resources<sup>3</sup>* |  |  |  |  |
| Partnered | 0.69 | 1 | 0.406 | 2726 |
| Social support | 1.75 | 2 | 0.173 | 2708 |
| Get along with others in household | 0.28 | 1 | 0.596 | 2708 |
| *Stressors<sup>4</sup>* |  |  |  |  |
| Additional stressful life events | 0.61 | 2 | 0.542 | 2708 |
| **Prior wave** |  |  |  |  |
| *Economic resources<sup>5</sup>* |  |  |  |  |
| Hardship | 0.86 | 2 | 0.425 | 2726 |
| Raise \$2000 in an emergency | 0.32 | 2 | 0.727 | 2445 |
| **History of ...** |  |  |  |  |
| *Mental health problems...* |  |  |  |  |
| Depressed mood<sup>6</sup> | 0.07 | 2 | 0.932 | 2659 |
| Anxiety<sup>6</sup> | 2.52 | 2 | 0.081 * | 2682 |
| *Economic resources* |  |  |  |  |
| Hardship<sup>6</sup> | 2.22 | 1 | 0.136 | 2726 |
| Raise \$2000 in an emergency<sup>6</sup> | 1.08 | 1 | 0.299 | 2676 |
| *Community resources* |  |  |  |  |
| Collective efficacy<sup>6</sup> | 0.60 | 2 | 0.551 | 2635 |
| *Parenting* |  |  |  |  |
| Harsh parenting<sup>6</sup> | 1.02 | 2 | 0.361 | 2726 |
| Warm parenting<sup>6</sup> | 0.00 | 1 | 0.999 | 2726 |

\* $p < 0.1$ ; \*\* $p < 0.05$ ; \*\*\* $p < 0.01$
<sup>1</sup>**Model 1 covariates:** fire exposure\*factor, flood, storm, cyclone, other disaster, previously exposed to disaster wave 4 to 8, sex, cohort, Indigenous background, neighborhood advantage/disadvantage, lived in regional/remote area, did not move in the past wave, born in overseas, state and territory.
<sup>2</sup>**Model 2 covariates:** Model 1 plus wave 9: employed and hardship
<sup>3</sup>**Model 3 covariates:** Model 2 plus wave 9 partnered, social support, get along others in household
<sup>4</sup>**Model 4 covariates:** Model 3 plus wave 9 stressful life events
<sup>5</sup>**Model 5 covariates:** Model 1 plus wave 1 to 8 hardship and raise \$2,000 in an emergency
<sup>6</sup>**Model 6 covariates:** Model 1 plus wave history independent variable

Table 3 shows the full regression results for the model with fire and history of anxiety predicting psychological distress, controlling for both demographic characteristics and previous exposure to disasters (waves 4 to 8). Results suggest that fire exposure only leads to higher levels of distress when Young People have some history of anxiety.

**Table 3:**
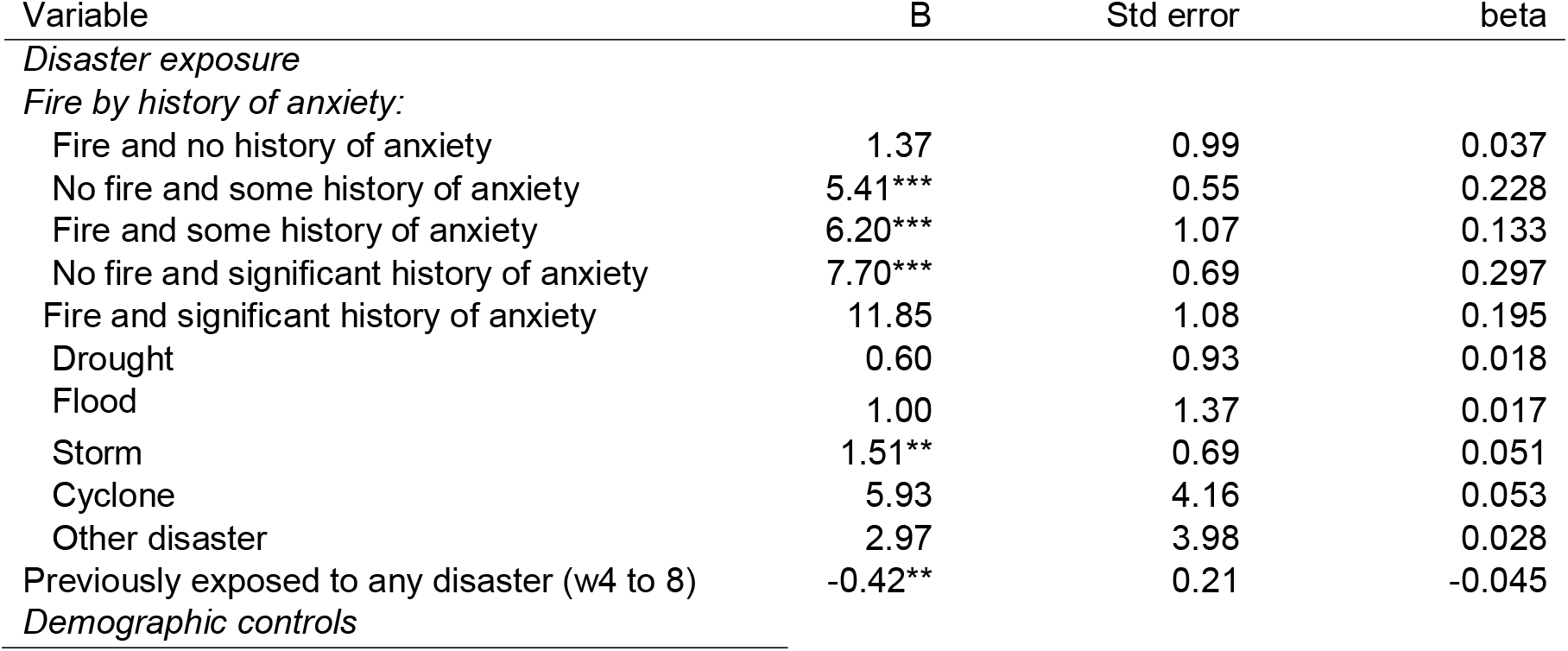
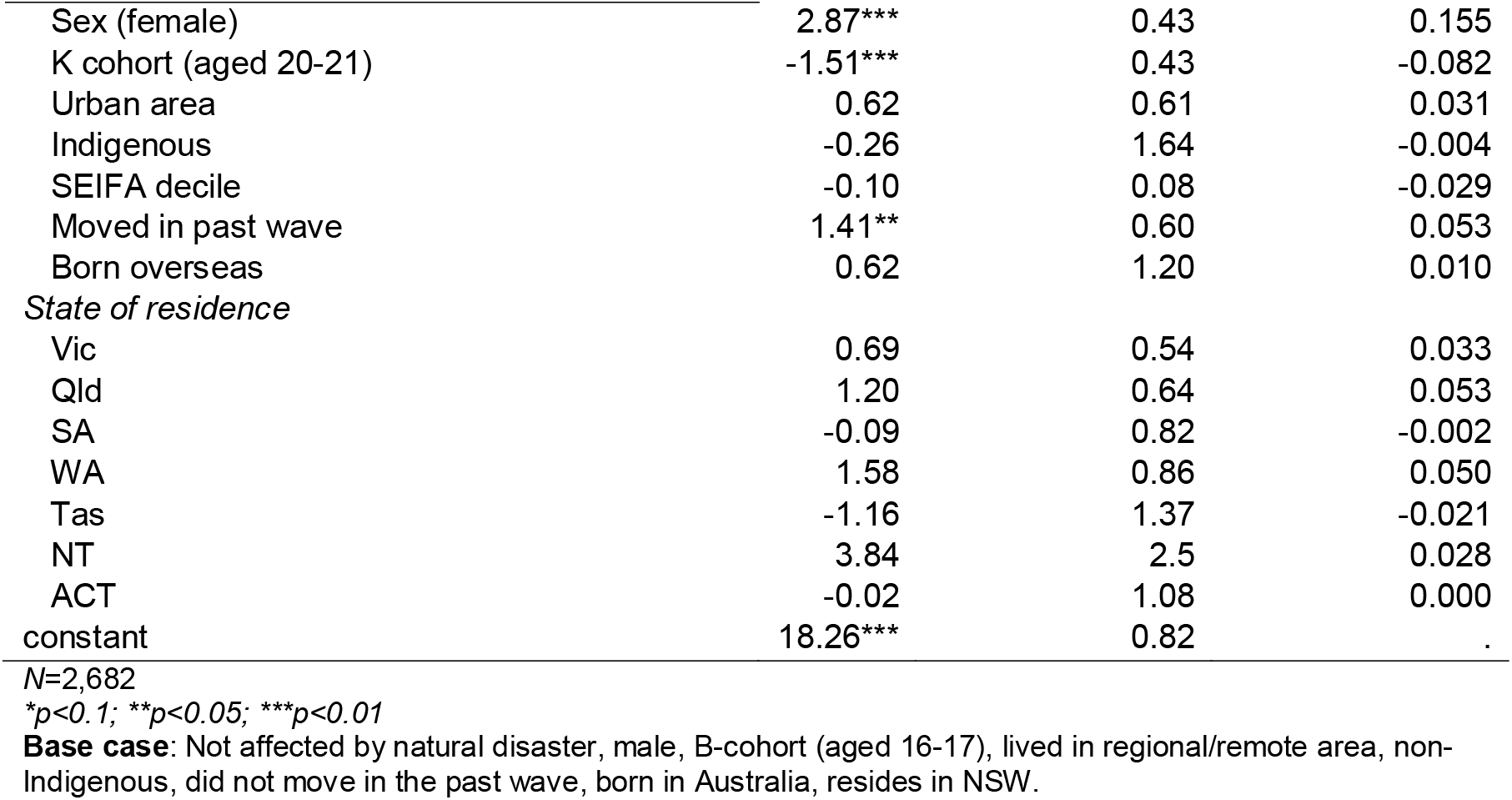
Self-report disaster exposure by history of high anxiety predicting psychological distress (K10), weighted linear regression.

| Variable | B | Std error | beta |
| --- | --- | --- | --- |
| *Disaster exposure* |  |  |  |
| *Fire by history of anxiety:* |  |  |  |
| Fire and no history of anxiety | 1.37 | 0.99 | 0.037 |
| No fire and some history of anxiety | 5.41*** | 0.55 | 0.228 |
| Fire and some history of anxiety | 6.20*** | 1.07 | 0.133 |
| No fire and significant history of anxiety | 7.70*** | 0.69 | 0.297 |
| Fire and significant history of anxiety | 11.85 | 1.08 | 0.195 |
| Drought | 0.60 | 0.93 | 0.018 |
| Flood | 1.00 | 1.37 | 0.017 |
| Storm | 1.51** | 0.69 | 0.051 |
| Cyclone | 5.93 | 4.16 | 0.053 |
| Other disaster | 2.97 | 3.98 | 0.028 |
| Previously exposed to any disaster (w4 to 8) | -0.42** | 0.21 | -0.045 |
| *Demographic controls* |  |  |  |
| Sex (female) | 2.87*** | 0.43 | 0.155 |
| K cohort (aged 20-21) | -1.51*** | 0.43 | -0.082 |
| Urban area | 0.62 | 0.61 | 0.031 |
| Indigenous | -0.26 | 1.64 | -0.004 |
| SEIFA decile | -0.10 | 0.08 | -0.029 |
| Moved in past wave | 1.41** | 0.60 | 0.053 |
| Born overseas | 0.62 | 1.20 | 0.010 |
| *State of residence* |  |  |  |
| Vic | 0.69 | 0.54 | 0.033 |
| Qld | 1.20 | 0.64 | 0.053 |
| SA | -0.09 | 0.82 | -0.002 |
| WA | 1.58 | 0.86 | 0.050 |
| Tas | -1.16 | 1.37 | -0.021 |
| NT | 3.84 | 2.5 | 0.028 |
| ACT | -0.02 | 1.08 | 0.000 |
| constant | 18.26*** | 0.82 | . |

N=2,682
\* $p < 0.1$ ; \*\* $p < 0.05$ ; \*\*\* $p < 0.01$
**Base case:** Not affected by natural disaster, male, B-cohort (aged 16-17), lived in regional/remote area, non-Indigenous, did not move in the past wave, born in Australia, resides in NSW.

Figure 1 shows that distress is significantly higher with some previous history of high anxiety and even greater again with a history of high anxiety in most waves. Those exposed to fire in the last 12 months and a history of high anxiety in most waves have significantly higher distress.

**Figure 1:**
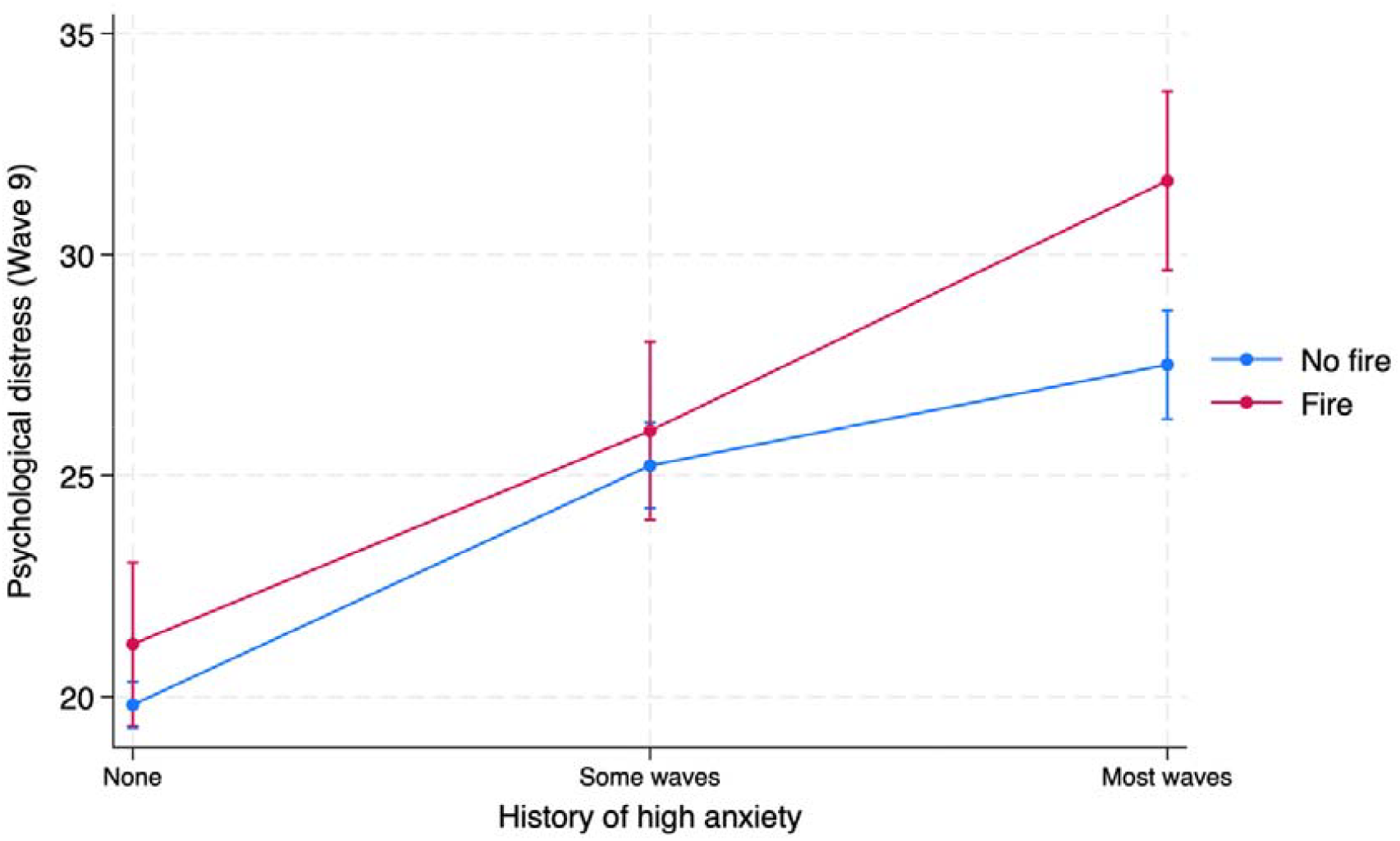
History of high anxiety by fire on psychological distress

## Discussion

In this study we systematically tested whether a range of economic, social and community resources protected against the negative impacts of bushfires on the psychological distress of young people. We also tested whether previous economic resources and longer-term economic resources over the previous 16 years protected against adverse mental health outcomes for youth. The protective effect of history of high parental warmth and low harsh parenting was also tested but the only statistically significant interaction with bushfire exposure was previous history of high anxiety. For those with no or some prior waves with high anxiety the 2019/20 fires was not associated with higher levels of distress but for those with a prior history for most waves, youth had significantly high levels of distress.

Despite testing a very large range of risk and protective factors identified in other studies as significant risk and protective factors in children and young people (Ma et al., 2022), we failed to find other significant moderators. This is unlikely to be due to a lack of statistical power as our study had one of the largest sample sizes of any examining risk and protective factors.

### Strengths and limitations

This study has several strengths in that it uses a large nationally representative sample. Based on Ma and colleagues’ review (2022) it is possibly the largest study to date in examining factors associated with mental health vulnerability and resilience when exposed to fires. Other strengths include detailed information about disaster exposures and controlling for disaster exposures in the previous decade along with other plausible covariates. We also comprehensively test a range of economic, social, and community resources. The use of longitudinal data on economic resources and parenting styles from early childhood is a strength as well as inclusion of previous history of mental health problems from age 12-13 years.

One limitation of the study is that for wave 9 data was being collected during the midst of a pandemic, but despite the huge variation in policy responses and outbreaks by state and territories (Edwards, et al., 2022) there were few significant differences by state and territory and regional indicators. However, this may have limited the capacity of economic, social, and community resources to protect against bushfires and further replication studies are needed to exclude this possibility.

## Conclusion

In this study we test whether economic, social, and community resources protect against detrimental mental health impacts of bushfires. While we do not find any evidence to support this notion, we do find that only those with a significant history of high anxiety have elevated distress levels. This evidence could be used in future to provide additional targeted support for those with prior history of mental health problems. Our findings underscore how robust the impacts of bushfires on the mental health of youth and that prior mental health history aside, being exposed to the Black Summer bushfires affected all youth irrespective of privilege or social address.

## Data Availability

Data is available from The Australian Data ArchivesDepartment of Social Services; Australian Institute of Family Studies; Australian Bureau of Statistics, 2021, Growing Up in Australia: Longitudinal Study of Australian Children (LSAC) Release 9.1 C1 (Waves 1-9), https://doi.org/10.26193/BAA3N6, ADA Dataverse, V6

## Notes

Conflict of interest: The authors have no real or perceived conflict of interest to declare.

### Competing Interest Statement

The authors have declared no competing interest.

### Funding Statement

Funding source: The work was conducted as part of an MHMRC Medical Research Future Fund grant (Grant ID: MRF1201335).

### Author Declarations

The study used ONLY openly available human data that were originally located at Australian Data Archives, https://doi.org/10.26193/BAA3N6

